# Psychiatric Hospitalization After Enrollment in Coordinated Specialty Care: Unexpected Gender and Age Related Disparities

**DOI:** 10.1101/2025.11.04.25339498

**Authors:** Jenifer L. Vohs, Sümeyra N. Tayfur, Fangyong Li, Zhiqian Song, Nicholas J. K. Breitborde, John Cahill, Serena Chaudhry, Maria Ferrara, Stephan Heckers, Audrey Satchivi, Steve Silverstein, Stephan F. Taylor, Ivy F. Tso, Ashley Weiss, Alan Breier, Vinod Srihari

## Abstract

**Background and hypotheses:** Hospitalization is common during first-episode psychosis (FEP) and is linked to functional decline, stigma, and healthcare burden. Coordinated Specialty Care (CSC) programs aim to reduce hospitalization and improve outcomes through early, multidisciplinary intervention. This study examined hospitalization outcomes and predictors among participants in the Academic Community Early Psychosis Intervention Network (AC-EPINET), a multisite CSC hub in the United States.

**Study design:** Participants with FEP (N = 701; mean age = 21.6 years, 64% male) were followed after CSC admission, with analyses restricted to the first 24 months. Primary outcomes included time to first hospitalization, number of hospitalizations, and length of stay (LOS). Kaplan-Meier survival and multivariable Cox regression examined predictors of time to first hospitalization, while negative binomial regression assessed hospitalization frequency and LOS.

**Study results:** Hospitalization rates declined after CSC enrollment. Females had shorter time to first hospitalization (HR = 2.96, 95% CI [1.24–7.10]) and more frequent admissions (IRR = 1.38, 95% CI [1.06–1.79]) than males. Younger age also predicted earlier (HR = 0.80, 95% CI [0.67–0.95]) and more frequent hospitalizations (IRR = 0.70 per 5 years, 95% CI [0.58–0.84]). Prior hospitalization predicted more admissions (IRR = 4.83, *p* < .0001) and longer LOS (RR = 10.72, *p* < .0001). Black/African American participants had longer LOS than White participants (RR = 1.67, *p* = .01).

**Conclusions:** While CSC reduces overall hospitalization risk, females, younger individuals, and those with prior admissions remain at elevated risk. These findings underscore the need for tailored strategies to mitigate disparities and optimize early psychosis care.

## 1. INTRODUCTION

Hospitalization is common in the early course of psychotic disorders and is associated with high personal and societal costs. Recent meta-analyses estimate that over half of individuals with first-episode psychosis (FEP) are hospitalized at their first presentation to care^1,2^. Further, examination of studies with an average seven-year follow-up period suggests that there is substantial risk of readmission, with a pooled average length of stay around 117 days^2^. Longer or repeated hospitalizations contribute to functional decline, stigma, and healthcare burden, making it critical to identify factors that predict hospitalization trajectories^2,3^. Prior studies have examined demographic and clinical predictors of hospitalization including sex^4,5^, age^4,5,6^, prior hospitalization^5,7^, poor social support^5^, duration of untreated psychosis (DUP)^4,7^, symptom severity^4,8^, substance misuse^7,9^, and poor medication adherence^7^, with mixed findings regarding which characteristics suggest highest risk .

Coordinated Specialty Care (CSC) programs have become the standard of care for FEP in the United States, emphasizing early engagement, family involvement, and recovery-oriented treatment^10^. To support discovery and improve outcomes, the National Institute of Mental Health launched the Early Psychosis Intervention Network (EPINET) in 2019, a national learning health system (LHS) encompassing eight regional hubs, over 100 CSC programs in 17 states, and a centralized data coordinating center^10^. The Academic Community Early Psychosis Intervention Network (AC-EPINET) is one such hub, comprising six CSC programs across three U.S. regions^11,12^. Leveraging harmonized measures and shared infrastructure, this network offers a unique opportunity to examine inpatient care outcomes across a large and diverse clinical sample. The present study focuses on longitudinal hospitalization outcomes after admission to CSC (i.e., time to first admission, number of episodes, and length of stay) among the AC-EPINET clinical sites and evaluates demographic and clinical predictors to identify subgroups at elevated risk.

## 2. METHODS

### 2.1. Sampling

AC-EPINET enrolled individuals with the following eligibility criteria: 1) aged between 12 and 35 years; 2) psychosis onset within the past 5 years; 3) affective or non-affective psychotic disorders confirmed by diagnostic interview and all other available sources of information (medical records, participant and family interviews); 4) outpatient status; 5) without a primary substance-induced psychosis disorder and 6) IQ>70 (determined by medical records and substantiated by testing when in doubt). A full description of AC-EPINET can be found elsewhere^11,12^.

### 2.2. Measures

All programs implemented the Core Assessment Battery (CAB), a standardized set of clinician-rated and self-report measures. The CAB assessed a range of domains including sociodemographic characteristics, pathways to care, psychiatric symptoms and diagnoses, psychosocial functioning and recovery, physical health, medication use, school and work participation, and substance use (https://nationalepinet.org/core-assessment-battery-cab/). Assessments are completed at program admission (baseline) and every six months thereafter for the duration of care received at the CSC. This analysis was restricted to variables collected during the 24 months following CSC admission.

The primary outcome for this analysis is hospitalization following CSC admission, characterized by: (i) time to first admission; (ii) number of hospitalization episodes, and (iii) length of hospital stay (LOS). Hospitalization data were obtained from self-report and, when available, corroborated by caretaker and medical record review. DUP (i.e., time from the onset of the first psychotic symptom to admission to CSC), and diagnosis were determined via clinical assessment and confirmed with any available additional information from caretakers and medical records. Cannabis use was confirmed via self-report. Symptoms were assessed with the Modified Colorado Symptom Index (MCSI), a 14-item self-report measure rated on a 0–4 scale (0 = not at all, 4 = daily), yielding a total score of 0–56 with higher scores indicating greater distress^13^. Functioning was assessed with the MIRECC-GAF^14^, which rates occupational and social domains on 1–100 scales (lower = greater impairment).

### 2.3. Statistical analysis

Time to first hospitalization was analyzed using Kaplan-Meier survival analysis stratified by sex. Further, multivariable Cox proportional hazards regression, with hazard ratios (HRs) and 95% confidence intervals (CIs) was used to adjust for sex, age, race/ethnicity, insurance status, education, DUP, and prior hospitalization. A separate model also adjusted for other covariates, including cannabis use, antipsychotic use, symptom severity, and functioning using a subset of data without missing values. To assess robustness of findings, sensitivity analyses were conducted. Excluding participants who were discharged from the CSC program yielded comparable results, and a supplementary competing-risk analysis treating known discharge reasons as a competing risk showed the same sex difference (see Supplementary File).

Among participants who were hospitalized, the number of hospitalization episodes was modeled using negative binomial regression, and results are reported as incidence rate ratios (IRRs) with 95% CIs. Length of stay (LOS) for hospitalization episodes was examined using generalized linear models with a log link and negative binomial distribution, reported as RRs with 95% CIs. Both models incorporated varying follow-up time among participants by including an offset in the model. Statistical significance was set at *p* < .05.

## 3. RESULTS

As shown in Table 1, the sample (N = 701) had a mean age of 21.6 years (SD = 3.9) and a median DUP of 7.7 months (IQR = 2.6 – 21.3). Most participants were male (64.1%) and identified as either White (42.5%) or Black/African American (37.7%). Ten percent were Hispanic/Latino. Over one-third had Medicaid insurance (36.7%), and nearly half (46.8%) had a high school education or less. Most (79.6%) had at least one hospitalization between psychosis onset and admission to CSC and 64.9% were receiving antipsychotic medication at baseline. The majority had a primary diagnosis of non-affective psychosis (75.5%). Symptom severity was mild (M = 22.3, SD = 15.2) with mild to moderate average occupational functioning (M = 53.2, SD = 24.0) and social functioning (M = 61.9, SD = 17.5).

**Table 1.**
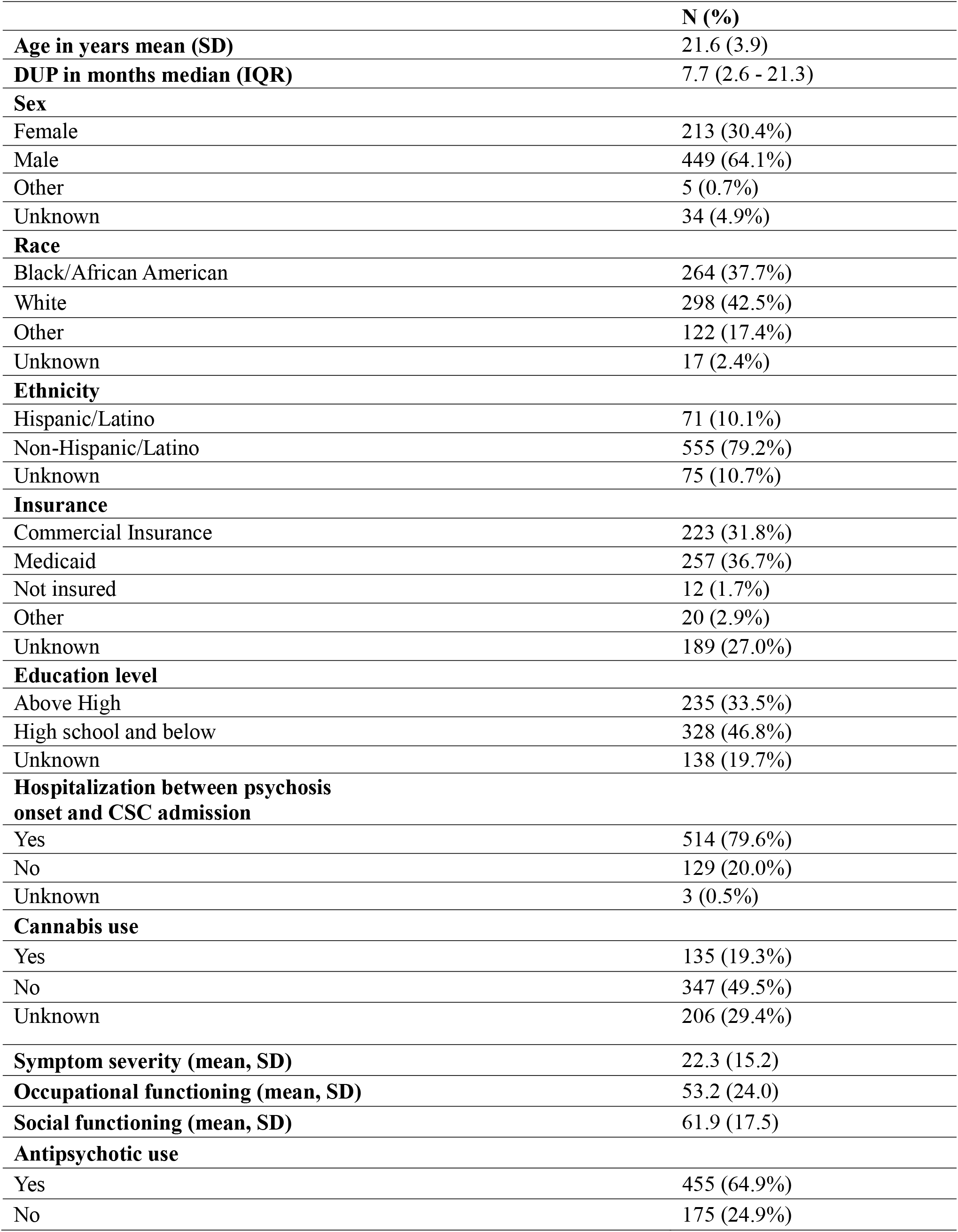
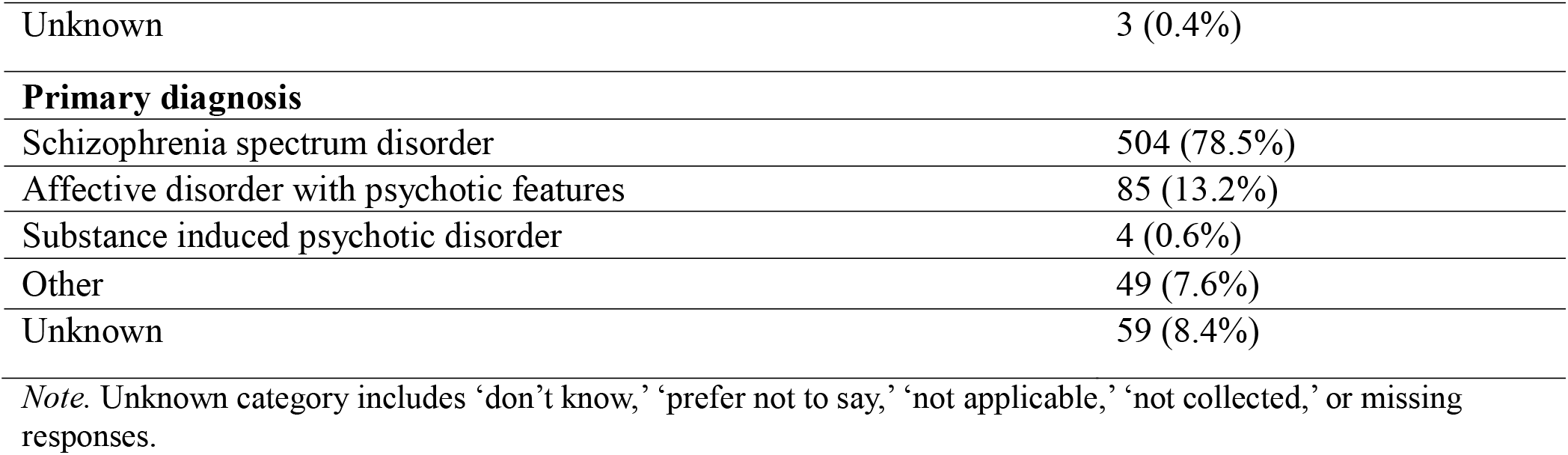
Baseline sample characteristics (N = 701)

|  | **N (%)** |
| --- | --- |
| **Age in years mean (SD)** | 21.6 (3.9) |
| **DUP in months median (IQR)** | 7.7 (2.6 - 21.3) |
| **Sex** |  |
| Female | 213 (30.4%) |
| Male | 449 (64.1%) |
| Other | 5 (0.7%) |
| Unknown | 34 (4.9%) |
| **Race** |  |
| Black/African American | 264 (37.7%) |
| White | 298 (42.5%) |
| Other | 122 (17.4%) |
| Unknown | 17 (2.4%) |
| **Ethnicity** |  |
| Hispanic/Latino | 71 (10.1%) |
| Non-Hispanic/Latino | 555 (79.2%) |
| Unknown | 75 (10.7%) |
| **Insurance** |  |
| Commercial Insurance | 223 (31.8%) |
| Medicaid | 257 (36.7%) |
| Not insured | 12 (1.7%) |
| Other | 20 (2.9%) |
| Unknown | 189 (27.0%) |
| **Education level** |  |
| Above High | 235 (33.5%) |
| High school and below | 328 (46.8%) |
| Unknown | 138 (19.7%) |
| **Hospitalization between psychosis onset and CSC admission** |  |
| Yes | 514 (79.6%) |
| No | 129 (20.0%) |
| Unknown | 3 (0.5%) |
| **Cannabis use** |  |
| Yes | 135 (19.3%) |
| No | 347 (49.5%) |
| Unknown | 206 (29.4%) |
| **Symptom severity (mean, SD)** | 22.3 (15.2) |
| **Occupational functioning (mean, SD)** | 53.2 (24.0) |
| **Social functioning (mean, SD)** | 61.9 (17.5) |
| **Antipsychotic use** |  |
| Yes | 455 (64.9%) |
| No | 175 (24.9%) |
| Unknown | 3 (0.4%) |
| **Primary diagnosis** |  |
| Schizophrenia spectrum disorder | 504 (78.5%) |
| Affective disorder with psychotic features | 85 (13.2%) |
| Substance induced psychotic disorder | 4 (0.6%) |
| Other | 49 (7.6%) |
| Unknown | 59 (8.4%) |
| *Note.* Unknown category includes ‘don’t know,’ ‘prefer not to say,’ ‘not applicable,’ ‘not collected,’ or missing responses. |  |

Figure 1 shows the change in hospitalization rates over the first two years after admission to coordinated specialty care. Hospitalization rates declined from baseline to 6 months and remained low throughout the 2-year follow-up period.

**Figure 1.**
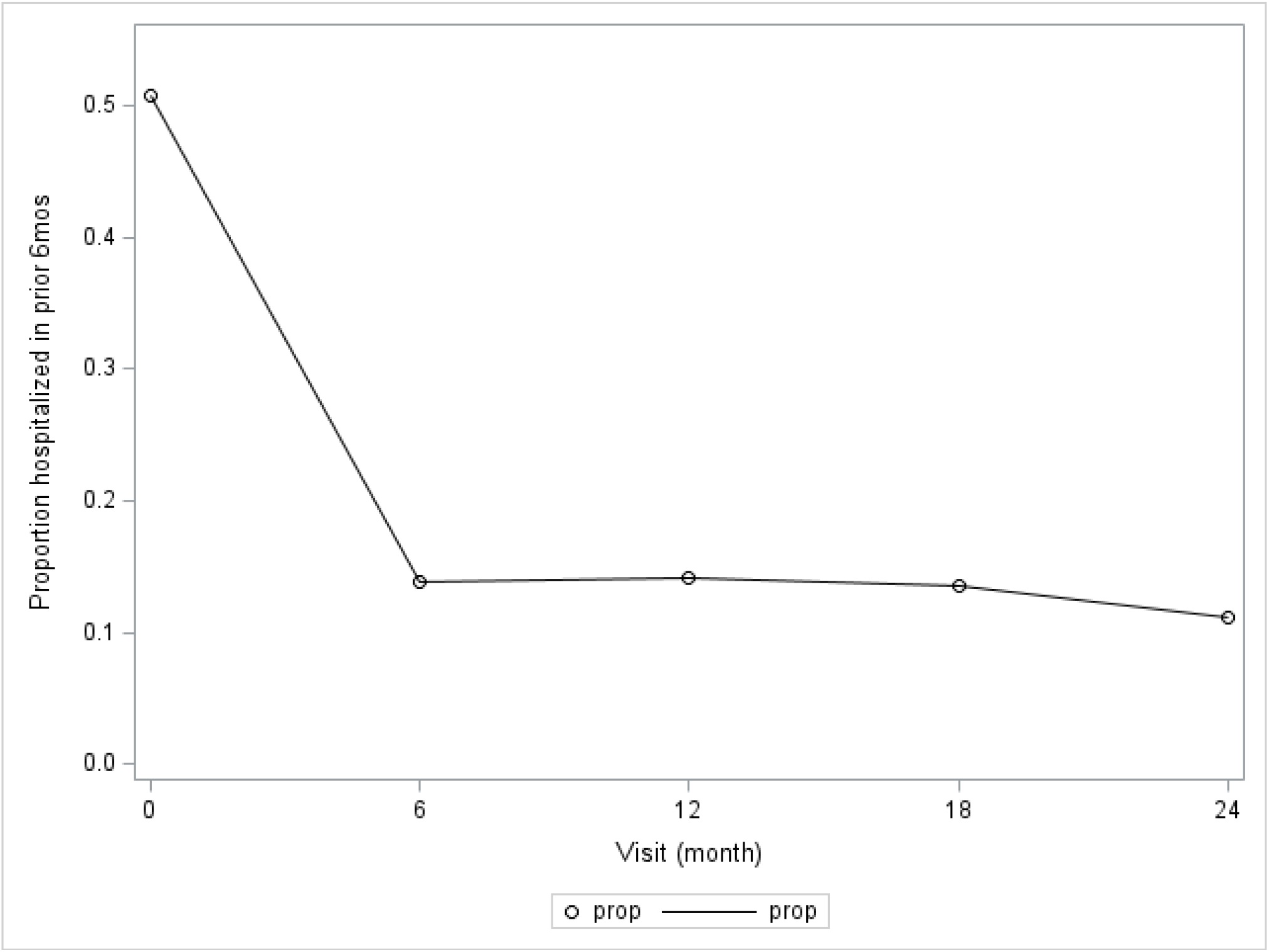
Overall change in hospitalization rates during the first 2 years of Coordinated Specialty Care (CSC)

As shown in Table 2, Cox proportional hazards regression analysis indicated that females had a significantly shorter time to first psychiatric hospitalization after admission to CSC compared to males (HR = 2.96, 95% CI [1.24–7.10], *p* = .02). Younger age was also associated with earlier hospitalization, with every one-year older age corresponding to a 19% lower hazard (HR = 0.80, 95% CI [0.67–0.95], *p* = .01). No significant differences were observed by race, ethnicity, insurance type, education level, DUP, or history of hospitalization in the 6 months prior to admission to CSC. Adjusting for baseline cannabis use, antipsychotic use, symptom severity, and both social and occupational functioning did not significantly change the results. Sensitivity analyses excluding discharged patients yielded comparable results supporting the robustness of the findings. Figure 2 shows the survival curves for time to first psychiatric hospitalization over the 2-year follow-up period. Based on the Kaplan-Meier curves, the hospitalization risk was 13.2% for females [CI: 7.5 – 22.5%] and 4.6% for males [CI: 2.3 – 9.2%] at 12-months.

**Table 1.**
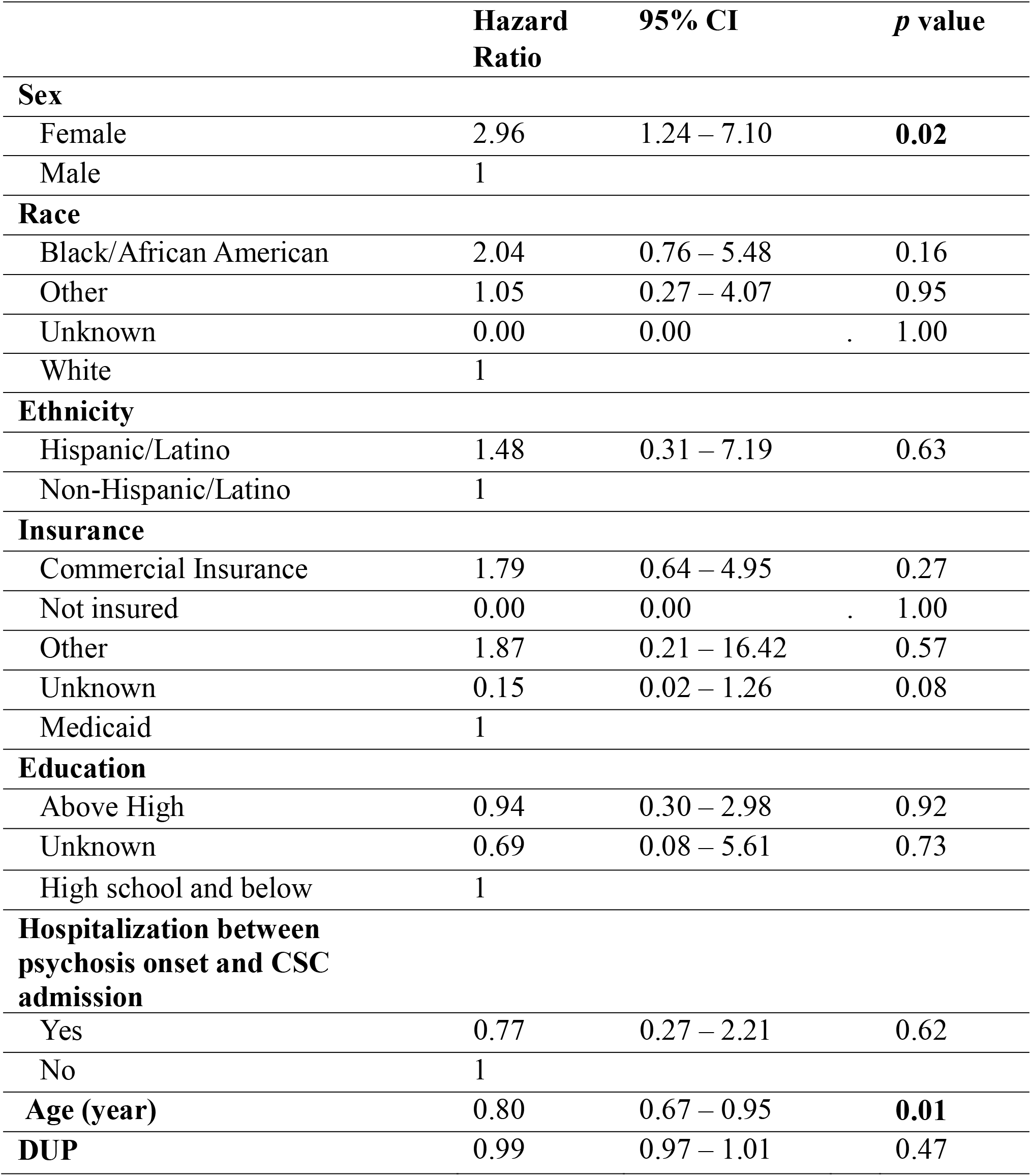
Cox regression model results.

**Figure 2.**
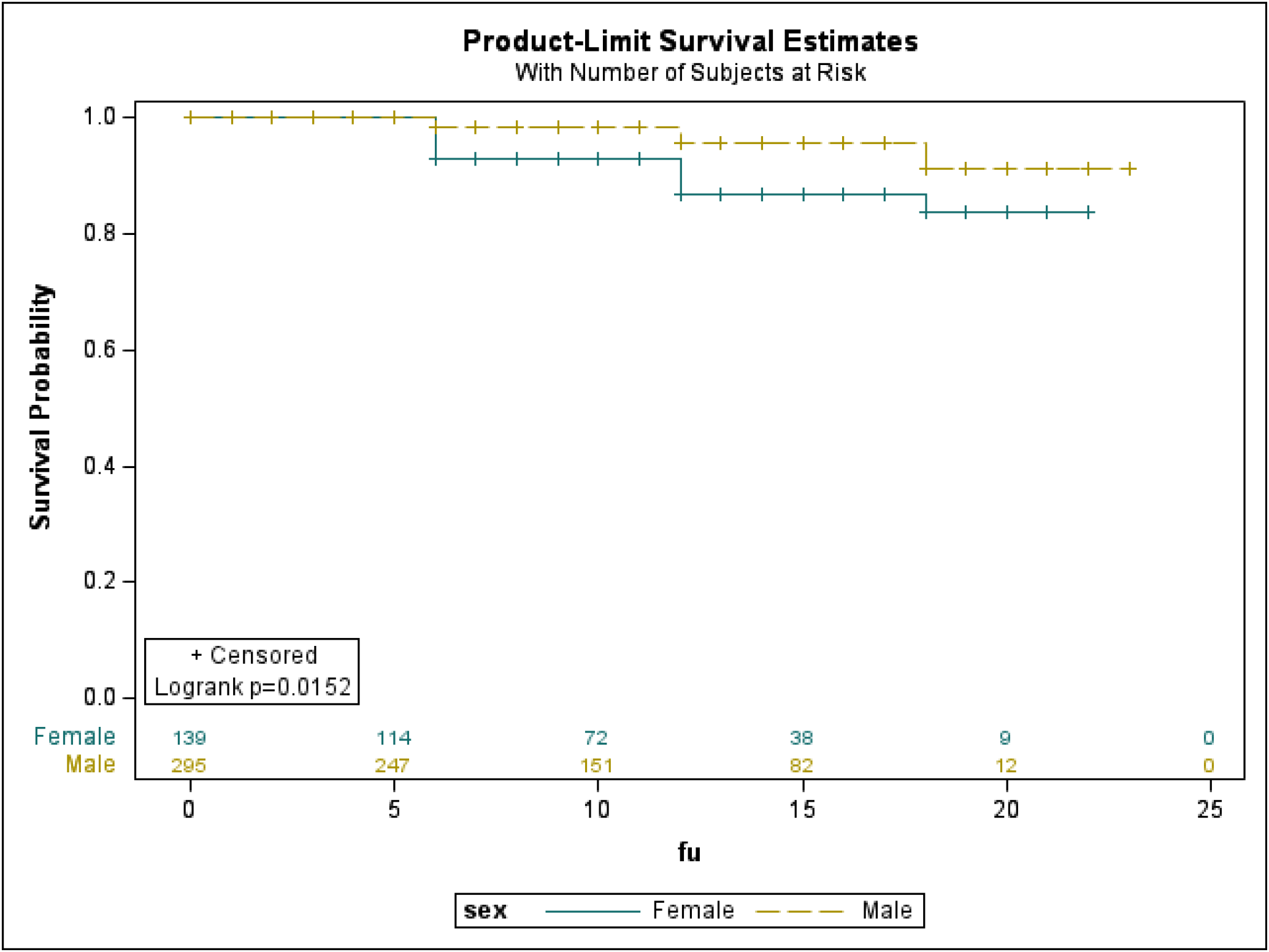
Time to first hospitalization in the first 2 years after admission to CSC

Figure 3a presents incidence rate ratios (IRRs) for the number of psychiatric hospitalization episodes among participants who were hospitalized during the two-year follow-up. Overall, episode rates were 51 per 100 person-years (95% CI: 35–75). Rates were 60 per 100 person-years for females (95% CI: 40–91) and 44 for males (95% CI: 30–65), and females had significantly higher rates (IRR = 1.38, 95% CI [1.06– 1.79], *p* = .02). Higher rates were observed among Black/African American participants, Hispanic participants, and those with more than a high school education. Hospitalization prior to CSC admission was strongly linked to more subsequent hospitalizations (IRR = 4.83, 95% CI [3.29–7.11], *p* < .0001). Longer DUP and older age were both associated with fewer hospitalization episodes. Specifically, each additional month of DUP was linked to a 1% decrease in the hospitalization episode rate (IRR = 0.99, 95% CI [0.98–1.00], *p* = .02), and every 5-year increase in age was associated with a 30% decrease (IRR = 0.70, 95% CI [0.58–0.84], *p* = .0002).

**Figure 3a.**
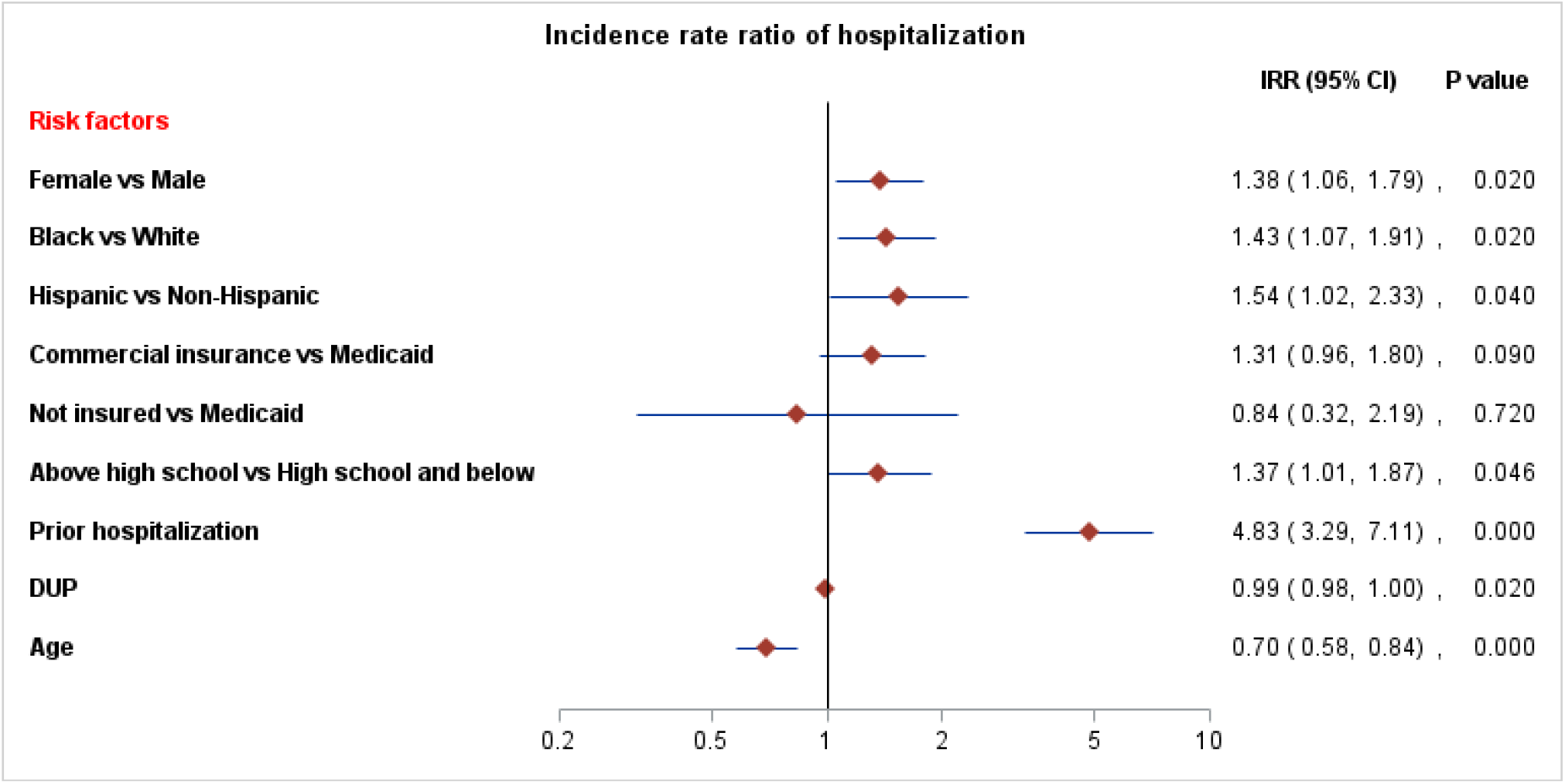
Number of psychiatric hospitalization episodes during the first two years after admission to CSC

**Figure 3b.**
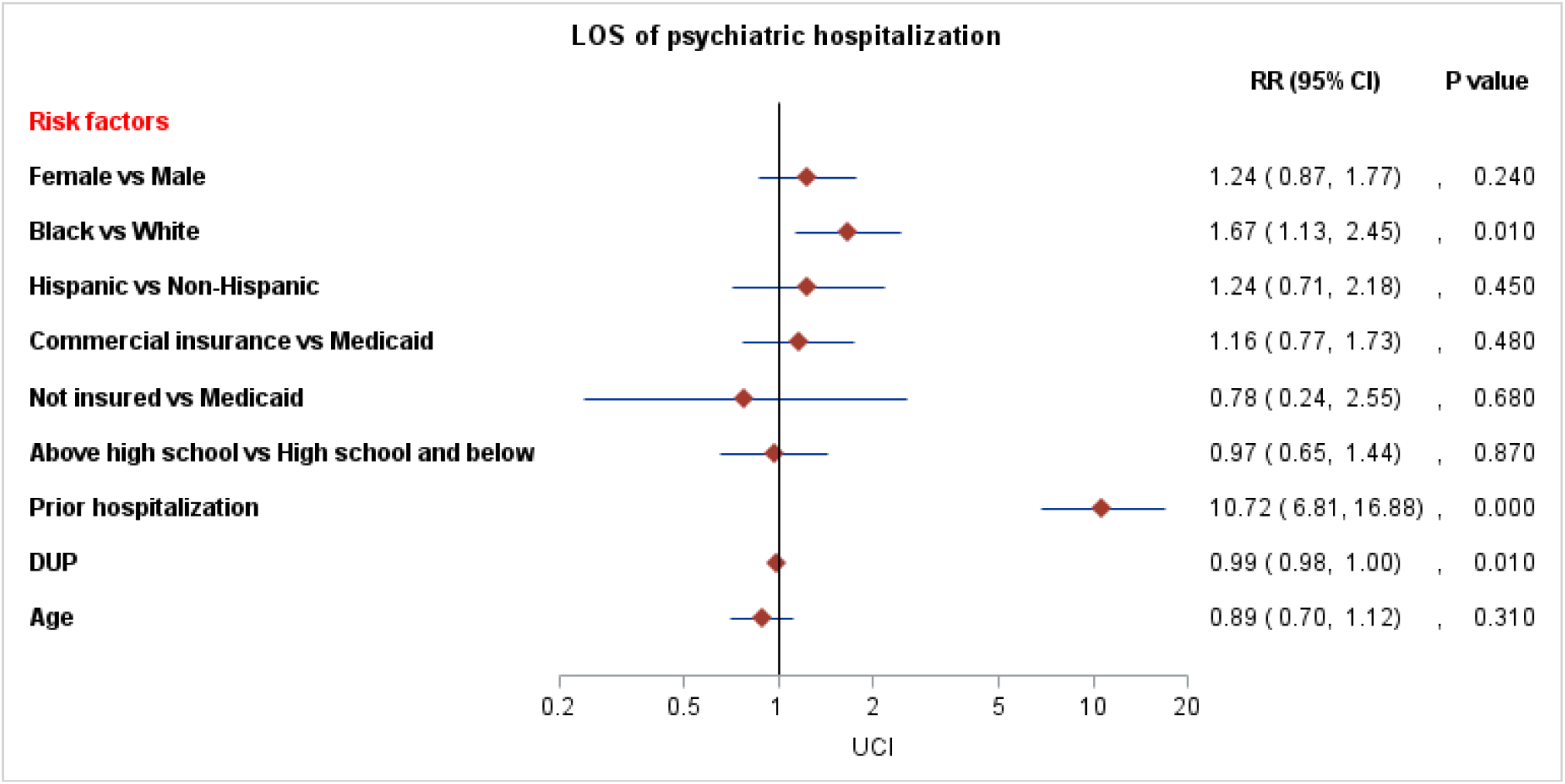
Length of stay (LOS) during the first two years after admission to CSC

Figure 3b presents regression results for the LOS among participants who were hospitalized during the two-year follow-up. Overall, participants spent average 3.7 days per year in the hospital (3.71, 95% CI: 2.30–5.98), with 4.1 days for females (95% CI: 2.5–7.0) and 3.3 days for males (95% CI: 2.0–5.5), although no significant difference was detectable (RR = 1.24, 95% CI [0.87–1.77], *p* = .24). However, Black/African American participants had significantly longer LOS than White participants (RR = 1.67, 95% CI [1.13–2.45], *p* = .01). There was no significant difference between Hispanic and non-Hispanic participants or between insurance types. Education level was not associated with differences in LOS. Hospitalization prior to CSC admission was strongly associated with longer stays (RR = 10.72, 95% CI [6.81–16.88], *p* < .0001). Each additional month of DUP corresponded to a 1% shorter LOS (RR = 0.99, 95% CI [0.98–1.00], *p* = .01). Age was not significantly associated with LOS (RR = 0.89, 95% CI [0.70– 1.12], *p* = .31).

## DISCUSSION

This multi-site study of individuals with FEP identified distinct patterns in hospitalization outcomes over the two years following admission to CSC programs. Although overall hospitalization rates declined, subgroup differences were evident. Females and younger participants consistently showed higher risk for earlier and more frequent hospitalizations, whereas length of stay (LOS) was associated with prior hospitalization history, DUP, race, but not with sex.

Sex differences at presentation may help explain the higher hospitalization rates observed among women. Females with FEP are more likely to present with affective symptoms, while males more often display negative symptoms^15,16^. In our data, women were also more likely to receive a diagnosis of affective psychosis (χ^2^ = 12.77, *p* < 0.001) and to report higher suicidal ideation at admission (χ^2^ = 6.12, *p* = .013). Suicidal behavior and depressive features, which are more common in women, are strong predictors of risk and may lower thresholds for hospitalization^17,18^. Interestingly, recent findings from an Italian early intervention cohort also reported unexpected sex differences: men showed lower symptom severity at 6 and 12 months and higher recovery rates at 12 months, although these differences converged by 24 months^19^. Longitudinal evidence similarly indicates that women’s early advantages such as better premorbid functioning, older age at onset, and higher recovery rates may diminish over time, resulting in outcomes more similar to men^20^. These patterns may also reflect different help-seeking behaviors, as women with FEP are more likely to seek care than men^21^. Another possibility may be differences in treatment practices. Several studies have shown that women with psychosis are less likely to receive clozapine or long-acting injectable antipsychotics, in part due to clinicians underestimating illness severity or assuming higher adherence to oral medications^19,22,23^. Such disparities in pharmacological management may compromise recovery and contribute to greater risk of relapse and hospitalization among women. These findings underscore the need for further investigation into how sex differences shape hospitalization trajectories in early psychosis.

Younger age also predicted earlier and more frequent admissions in the current study, consistent with prior evidence linking younger onset to poorer trajectories and increased service use^2,6^, possibly mediated by greater family involvement in help-seeking and admission decisions. Importantly, neither sex nor age was associated with LOS, suggesting their influence lies primarily in the likelihood of admission rather than inpatient course once admitted. Future work utilizing the larger, nationwide datasets including other EPINET sites and claims databases could further test the generalizability of these findings across diverse CSC programs.

As expected, prior hospitalization was the strongest predictor of both number of episodes and LOS. Individuals with hospitalization history were at elevated risk of more frequent episodes and longer stays. This finding is consistent with longitudinal research identifying prior hospitalization as a robust marker of severity and relapse risk^7,8^. We also found evidence of disparities by race: Black/African American participants had significantly longer LOS compared to White participants, consistent with longstanding concerns about systemic inequities in psychiatric care^2,24^. These findings may reflect differences in pathways to care, cultural stigma, or systemic biases in hospitalization thresholds^2,25^. Addressing these inequities requires both structural changes and culturally responsive engagement strategies within CSC programs.

The association between DUP and hospitalization is complex. Longer DUP was linked to fewer admissions and shorter LOS in this cohort. A previous study of a sample with shorter overall delays to care reported an association between longer DUP and increased hospitalizations within the first year of CSC^26^. A meta-analysis of a more heterogenous collection of FEP cohorts and longer follow-up periods reported no significant relationship between DUP and hospital admissions or LOS^27^. More recently, Gannon et al. (2024) also found that longer DUP predicted fewer hospital admissions, consistent with our findings^4^. One explanation for these inconsistent findings is the varied proportion of patients with insidious illness course. These individuals have subtler and more negative symptoms that are less socially disruptive and thus less likely to trigger hospitalization. Separately, Robinson et al. (2019) have argued that studies that include more participants with already short DUP fail to detect associations with hospitalization due to a floor effect (i.e., further reductions in delay can do little to measurably reduce hospitalization risk)^7^. Also, DUP was not assessed with standardized tools but relied on clinician impression which may have underestimated or compressed variability in estimates of delay. Prolonged DUP remains one of the most consistent predictors of worse long-term clinical and functional outcomes^27^ and this has motivated efforts to reduce delays to care. These findings suggest that such early detection efforts, if successful at shortening DUP to CSC, may have effects on hospitalization that vary based on sample characteristics, treatment models, and duration of follow-up.

Several limitations should be considered when interpreting these findings. Clinical variables previously associated with hospitalization in FEP, such as cannabis use, baseline symptom severity, and functional impairment, were not significant predictors in our models, which was unexpected given prior research linking these factors to relapse and hospitalization risk^2,8^. This may have been a Type II error caused by missingness across several clinical measures (e.g., symptom severity missing: 34.7%). Additionally, DUP was not measured with a psychometrically valid tool and may have been subject to both systematic and random error across sites. The unexpected finding of greater hospitalization burden among women should thus be interpreted cautiously. Our sensitivity analysis did not show significant sex differences in disengagement from care, reducing concern about attrition bias. However, hospitalization data were drawn from self-report, caregiver input, and medical records, and some degree of sex-related reporting bias cannot be ruled out given potential differences in disclosure or caregiver involvement. Moreover, the dataset did not distinguish voluntary from involuntary admissions or capture whether hospitalizations were driven by suicidality versus psychotic relapse, factors that may be more common among women and contribute to higher admission rates. Finally, practice variation across sites in thresholds for hospitalization could interact with sex in ways not fully captured here.

## CONCLUSIONS

Engagement in CSC was associated with reduced hospitalization risk over the two-year follow-up, though a significant proportion of participants continued to experience admissions. Females and younger participants were at disproportionate risk for earlier and more frequent hospitalizations. Prior hospitalization and racial disparities were important determinants of LOS. These findings highlight opportunities to refine early intervention strategies for high-risk subgroups entering CSC.

## Data Availability

All data produced are available online at ENDCC (EPINET Data Coordinating Center).

## Conflict of Interest

The authors report no conflicts of interest.

## Funding

This work was supported by the National Institute of Mental Health (NIMH; R01MH120588).

## Notes

### Competing Interest Statement

The authors have declared no competing interest.

### Funding Statement

The ENDCC is supported by the National Institute of Mental Health under award number 5U24MH120591.

### Author Declarations

All participating clinics in the EPINET initiative operate under institutional review board or equivalent ethics committee approval, and data shared with the national coordinating centre are de-identified and governed by data use agreements.

