## Supplemental File for "Psychiatric Hospitalization After Enrollment in Coordinated Specialty Care: Unexpected Gender and Age Related Disparities"

**Appendix A. Discharge Categories**

| **Code** | **Discharge Categories** |
| --- | --- |
| 1 | Completed program, graduated, or services no longer indicated due to improvement |
| 5 | Whereabouts unknown, team unable to contact client |
| 6 | Incarcerated |
| 7 | Admitted to a state hospital |
| 9 | Other |
| 10 | Terminated, refused or declined services |
| 11 | Client does not display signs/symptoms warranting covered diagnosis/impairment |
| 12 | Reached limit for length of allowable stay |
| 13 | Pursuing a positive opportunity (e.g., school, employment, training) |
| 14 | Admitted to a residential program |
| 15 | Transferred services to provider outside CSC program (not state hospital/res.) |
| 16 | Moved out of service area (other reasons) |
| 17 | Deceased (by suicide) |
| 18 | Deceased (by other means) |
| 7777 | Not Applicable |
| 8888 | Not Collected |
| 9999 | Missing |

**Figure.** Sensitivity analyses censoring participants with unclear discharge reasons (codes 5, 9, 7777, 8888, 9999) versus those with defined reasons (cumulative incidence function)

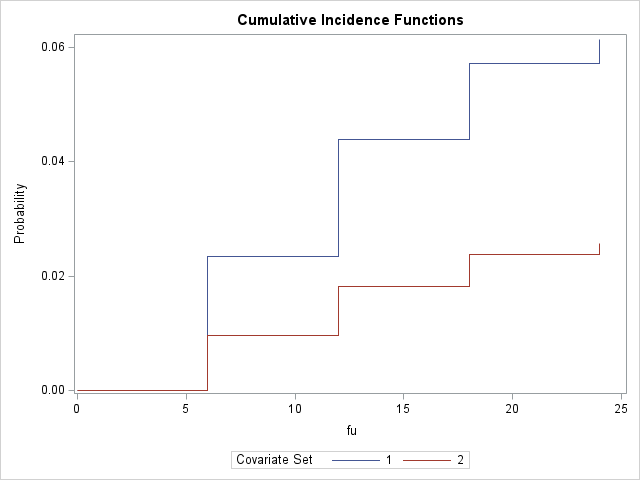
*Note.* Adjusted for race, ethnicity, insurance status, education level, prior hospitalization, age, DUP.
